# *MALAT1* Expression is Associated with Aggressive Behavior in Indolent B-Cell Neoplasms

**DOI:** 10.1101/2023.02.15.23285907

**Authors:** Elena María Fernández-Garnacho, Ferran Nadeu, Silvia Martín, Pablo Mozas, Andrea Rivero, Julio Delgado, Eva Giné, Armando López-Guillermo, Martí Duran-Ferrer, Itziar Salaverria, Cristina López, Sílvia Beà, Santiago Demajo, Pedro Jares, Xose S Puente, José Ignacio Martín-Subero, Elias Campo, Lluís Hernández

## Abstract

*MALAT1* is a long non-coding RNA with oncogenic roles in cancer but poorly studied in indolent B-cell neoplasms. Here, *MALAT1* expression was analyzed using RNA-seq, microarrays or qRT-PCR in primary samples from various clinico-biological subtypes of chronic lymphocytic leukemia (CLL, n=266) and follicular lymphoma (FL, n=61). In peripheral blood (PB) CLL samples, high *MALAT1* expression was associated with a significantly shorter time to treatment, independently from other known prognostic factors, such as IGHV mutational status. Coding genes whose expression levels were associated with *MALAT1* in CLL were predominantly related to oncogenic pathways stimulated in the lymph node (LN) microenvironment. Further analysis of *MALAT1* expression by microarrays in paired CLL samples from PB/LN showed that its levels were maintained between both anatomical compartments, supporting that the clinical value of *MALAT1* expression found in PB is mirroring expression differences already present in LN. Similarly, high *MALAT1* expression in FL predicted for a shorter progression-free survival, and its correlated expressed genes were associated with pathways promoting FL pathogenesis. In summary, *MALAT1* expression is related to pathophysiology and clinical behavior of indolent B-cell neoplasms. Particularly in CLL its levels could be a surrogate marker of the microenvironment stimulation and may contribute to refine the clinical management of these patients.

## Introduction

Long non-coding RNAs (lncRNAs) regulate the expression of protein-coding genes and are increasingly described as key players in physiological and pathological conditions, most remarkably in cancer^1,2^. One of the lncRNAs most frequently related to oncogenesis is *MALAT1*, which has been implicated in the regulation of key cellular pathways such as MAPK/ERK, PI3K/AKT, WNT/B-catenin, and NF-kB^3^ and, as a consequence, involved in many cancer-associated processes such as cell proliferation, migration, invasion, apoptosis and angiogenesis^4^. There are also several studies supporting *MALAT1* expression as a clinical biomarker mainly associated with a poor prognosis in solid tumors^3^, although in some neoplasms such as diffuse large B-cell lymphoma (DLBCL), colorectal and breast cancer, high levels of *MALAT1* have been linked to a favorable outcome^5–8^.

In B-cell non-Hodgkin lymphomas (B-NHL), we and others have previously shown that lncRNA deregulation is associated with immune cell-related functions and cell proliferation control^9,10^. In the case of *MALAT1*, its expression has been shown to be upregulated in some lymphoid neoplasms such as DLBCL^11^, chronic lymphocytic leukemia (CLL)^12^, and mantle cell lymphoma (MCL)^13^. These results suggest that *MALAT1* expression could be associated with poor prognosis even in indolent B-NHL, but most of these studies include relatively few cases and the possible clinical relevance of *MALAT1* expression in these lymphoid neoplasms is still not well known. In this study, we have performed a detailed characterization of the biological and clinical impact of *MALAT1* deregulation in two types of indolent B cell lymphomas with different underlying pathobiological mechanisms, namely CLL and follicular lymphoma (FL).

## Material and methods

### Description of the transcriptional data and patient cohorts

This study exploited previously published genome wide transcriptional data from CLL and FL. Regarding CLL, we reanalyzed our RNA-seq data from peripheral blood (PB) samples obtained from 266 patients, and 25 additional monoclonal B cell lymphocytosis donors (MBL) of the Spanish ICGC consortium with full clinical annotations^14^. We have also analyzed the microarray data from the GSE21029 GEO dataset, including 17 patients with paired PB and lymph node (LN) samples and 7 additional samples from PB^15^. For the analysis of *MALAT1* in FL, we extracted RNA from formalin-fixed paraffin-embedded (FFPE) LN samples of 61 FL patients (stages 1 to 3A) from the Hematopathology collection of the Biobank of the Hospital Clínic de Barcelona-IDIBAPS (Spain). In addition, microarray data generated from sorted FL neoplastic cells of 23 patients were obtained from the GSE107367 GEO dataset^16^.

### RNA extraction and qRT-PCR

For the 61 FFPE FL samples, 3-10 cuts of 10*μ*m each were used per sample for RNA extraction using Allprep DNA/RNA FFPE kit (Qiagen). RNA integrity was analyzed with TS4200 using DV200 index as recommended by the manufacturer (Agilent). In this way, samples with DV200 less than 35% were excluded to avoid excessive degraded material. Reverse transcriptase reaction was performed using High-Capacity Reverse transcription kit (Applied Biosystems) with an RNA input of 90 ng.

*MALAT1* expression was analyzed by qRT-PCR using a short amplicon specifically designed for FFPE samples. *MALAT1* expression levels were normalized using three reference genes (*ACTB, GAPDH* and *YWHAZ*) which have been previously used in B-cell lymphomas, including FL samples^17,18^. Primer quantities were optimized for each amplicon to reach a high efficiency (range: 90-96%) and their sequences were: 5’-CCCCTTCCCTAGGGGATTTCA-3’ (*MALAT1* forward), 5’-AAGCCCACAGGAACAAGTCC-3’ (*MALAT1* reverse), 5’-CCAACCGCGAGAAGATGAC-3’(ACTB forward), 5’-TAGCACAGCCTGGATAGCAA-3’ (*ACTB* reverse), 5’-AGGTGAAGGTCGGAGTCA-3’(*GAPDH* forward), 5’ CAACAATATCCACTTTACCAGAGTTAA-3’ (*GAPDH* reverse), 5’-CAAAGACAGCACGCTAATAATGCA-3’ (*YWHAZ* forward), and 5’-TCAGCTTCGTCTCCTTGGGTA-3’ (*YWHAZ* reverse). Amplification reactions were carried out using PowerUp™ SYBR™ Green Master Mix (Applied Biosystems, Foster City, CA) following supplier’s recommendations in a Step One Plus thermocycler (Applied Biosystems). Transcript expression relative quantification was performed referred to a calibrator sample of universal human reference RNA (Invitrogen).

### Bioinformatic analyses and statistics

Microarray data were normalized with RMA-based methodology and used to analyze the expression levels of *MALAT1* transcript. Although the expression microarrays were enriched in probe sets for coding genes, several probe sets that hybridize exclusively to the *MALAT1* transcript were initially included. The specificity of the probe sets was confirmed in the Affymetrix database (NettAffx™). Only one probe set was excluded (223579_s_at) at this stage. The correlations among the different probe sets were checked by strand and another probe set was excluded as an outlier (224559_at). Finally, the mean values of the remaining probe sets were retained as reliable for measuring the expression levels of *MALAT1* (probe sets 1558678_s_at, 223940_x_at, 224558_s_at, 224567_x_at, 224568_x_at and 226675_s_at).

To identify potential target genes, ranked lists of coding genes according to the degree of Pearson’s correlation to *MALAT1* were obtained for the different indolent B-cell lymphomas under study using either RNA-seq or microarray data. Only those statistically significant correlations after correction for multiple comparisons (adjusted p-value<0.05) were considered for the downstream analyses. We separately analyzed positive and negative correlations of coding genes with *MALAT1* using the webtool Metascape^19^. The Metascape output of enriched (Reactome) pathways were obtained as tables, and summary plots depicting relevant pathways with very high statistical significance (q<0.05) were prepared with Graphpad Prism v7. Furthermore, we used Gene Set Enrichment Analysis (GSEA v3.0 from http://www.gsea-msigdb.org) to perform additional pathway enrichment analyses in CLL using the C2 curated gene signatures from MSigDB/GSEA website and several specific signatures previously related to differences between LN and PB in CLL ^20^. Only statistically significant signatures with NES higher than 2 as absolute value were considered as potentially relevant.

In the clinical association studies, optimal cut-off points for *MALAT1* expression groups were obtained using the *maxstat* algorithm which optimized the log-rank statistics (*maxstat* package, R Software, Vienna). Cumulative incidence and Kaplan-Meier curves, scatterplots, and box plot graphs were generated using both the R environment and GraphPad Prism v7. Univariate and multivariate Fine-Gray regression models considering *MALAT1* as a continuous variable (as a more stringent statistical approach) were used for measuring its impact on time to treatment (TTT). Univariate analysis of *MALAT1* as a continuous variable was performed with the Cox regression model for overall survival (OS). Both calculations were performed with R v.4.0.3. Student’s t-test was used to evaluate the differences in the mean expression of *MALAT1* among subtypes and molecular factors with previously described prognostic value in CLL. Paired t-test was used to evaluate differences in *MALAT1* expression between paired samples of CLL in PB and LN^15^. Linear regression on scatter plots was representing the correlation between *MALAT1* expression and the epiCMIT score (epigenetically-determined Cumulative MIToses)^21^.

## Results

### MALAT1 expression in CLL

We initially evaluated the expression levels of *MALAT1* in CLL and MBL patients. *MALAT1* expression was significantly higher in CLL than in MBL (P<0.001) (Fig. 1A). We also stratified *MALAT1* levels according to the IGHV mutational status (mutated vs non-mutated) and epigenetic subtypes (naïve-like, memory-like and intermediate) and no significant differences were observed among the different groups (Fig. 1A).

**Figure 1.**
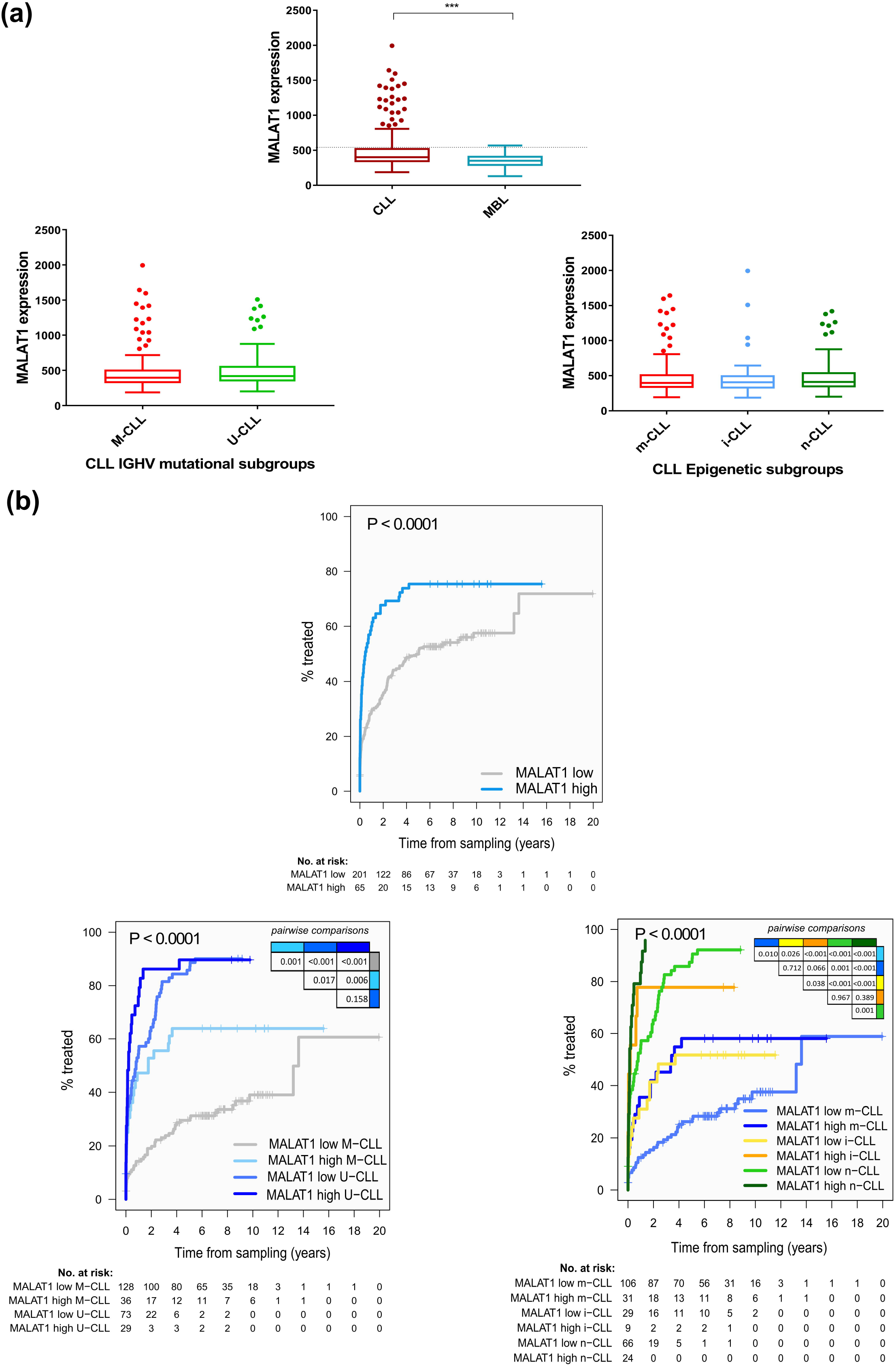
*MALAT1* expression levels in MBL, CLL molecular subgroups, and relationship to time to treatment (TTT). **(a)** *MALAT1* expression was higher in CLL patients than in MBL (P<0.001). Dotted line indicates the threshold of *MALAT1* that determines differences in outcome. No significant differences were observed in *MALAT1* expression between CLL subgroups defined by IGHV mutational status (M-CLL versus U-CLL) (bottom left panel) or epigenetic subtypes (n-CLL: naïve-like, m-CLL: memory-like and i-CLL: intermediate) (bottom right panel). **(b)** CLL patients with high *MALAT1* expression showed significantly shorter TTT compared to those with low levels in the global cohort (top panel), and almost every one of the different CLL subgroups related to the IGHV mutational status (bottom left) and epigenetic subtypes (bottom right).

We next evaluated whether *MALAT1* expression was associated with the clinical behavior of the tumor. Using the *maxstat* algorithm, we segregated CLL patients into high and low expression groups (Fig. S1A). Patients with high *MALAT1* expression (N=65) had a significantly shorter time to treatment (TTT) than the *MALAT1*-low group (N=201) (p<0.0001) (Fig. 1B). This finding was also confirmed in the subset of patients clinically classified as Binet A (N=239, 57 with high and 182 low *MALAT1* levels) (P=0.0003) (Fig. S2A). On the contrary, *MALAT1* levels were not related to the overall survival (OS) of the patients (Fig. S1B and Fig. S2B). The adverse impact of high *MALAT1* expression on TTT was further confirmed in IGHV-mutated CLL and in the three epigenetic CLL subtypes (Figure 1B). Similar findings were observed when the analyses were restricted to patients with Binet A CLL (Fig. S2C-S2D). To evaluate the independent prognostic impact of *MALAT1*, we performed multivariate regression models. First, we checked that *MALAT1* as a continuous isolated variable also had significant prognostic value for TTT (HR=1.32; 95%CI: 1.18-1.48); p<0.0001) but not for OS (HR = 1.05; 95%CI: 0.89-1.24; p=0.561). Next, the multivariate analyses confirmed that *MALAT1* expression also had an independent prognostic value for TTT considering Binet stage and the IGHV mutational status (P=0.0004) and Binet stage and the epigenetic subgroups (P<0.0001) (Fig. 2A). We also evaluated the possible association of *MALAT1* levels with other molecular factors previously shown to have prognostic value in CLL, such as mutations in driver genes, number of chromosomal aberrations^14,22^, IGLV3-21 variant/R110 mutation^23^, or the DNA methylation-based epiCMIT score related to the proliferative history of the tumor cells^21^. *MALAT1* expression levels were not related to any of these variables (Fig. S3 and Fig.S4). Overall, these data indicate that *MALAT1* expression has a prognostic value for TTT in CLL irrespective of other known genetic and epigenetic prognostic parameters.

**Figure 2.**
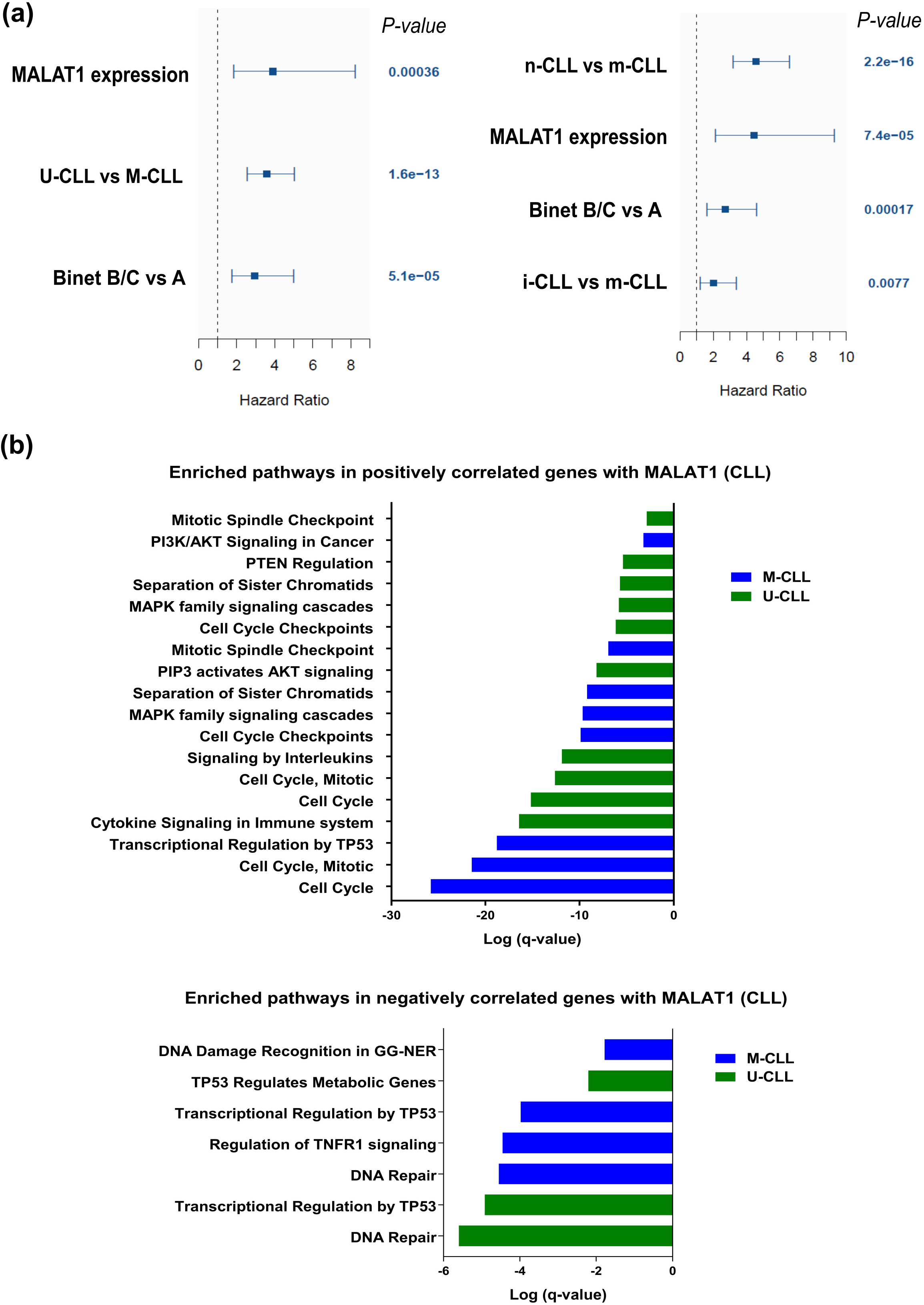
*MALAT1* expression is an independent prognostic factor and is related to relevant pathways in CLL. **(a)** Plot for multivariate model analyses, where *MALAT1* expression as a continuous variable shows its independent prognostic value considered together with Binet stage and IGHV mutational status (left panel) or epigenetic subtypes (right panel). For epigenetic groups, m-CLL was taken as a reference. **(b)** Summary of most relevant significant pathway enrichments found using Metascape tool regarding coding genes positively (top panel) and negatively (bottom panel) correlated with *MALAT1* expression in CLL subtypes defined by IGHV mutational status. Only statistically significant Reactome pathways after multiple comparison correction are shown for compact representation.

To determine whether *MALAT1* deregulation could be related to genetic alterations, we analyzed the gene mutational status in the whole-genome sequences of 150 cases from the ICGC Consortium^14^. Only five CLL and one MBL revealed mutations in the *MALAT1* locus and were not related to the expression of the gene (Table S1A). No copy number alterations affecting 11q13, where *MALAT1* is located, were found, whereas only 2 CLL cases had copy number neutral loss of heterozygosity. None of these alterations were related to the expression of the gene (Table S1B).

To evaluate the possible functional implications of *MALAT1* expression we searched for coding genes that were significantly correlated, either positively or negatively, with *MALAT1* in the different CLL subgroups (Table S2) and we subsequently performed pathway enrichment analyses. Among the positively correlated genes, the significantly selected pathways were remarkably associated with functions related to activation, proliferation, and survival of CLL cells in the LN microenvironment (Fig. 2B, Fig. S5, Table S3 and Table S4). These pathways included general cell proliferation-related functions such as signaling by PI3K/AKT, MAPK, IL10 and IL4, among others^24–29^. As these results pointed to a possible influence of microenvironment stimuli in *MALAT1* expression we used GSEA to analyze CLL specific signatures and finding, both in U-CLL and M-CLL, significant positive enrichments in a signature of genes previously described as significantly upregulated in LN vs. PB (Fig. 3B)^20^. Finally, we also analyzed previously published microarray data of paired LN and PB CLL samples^15^, where we found that *MALAT1* levels did not change significantly between LN and PB (P=0.097) (Fig. 3C). All these results suggest that the relationship between *MALAT1* expression and shorter TTT that we have observed in PB samples could represent a surrogate biomarker for the degree of stimulation and proliferation of CLL cells in the LN.

**Figure 3.**
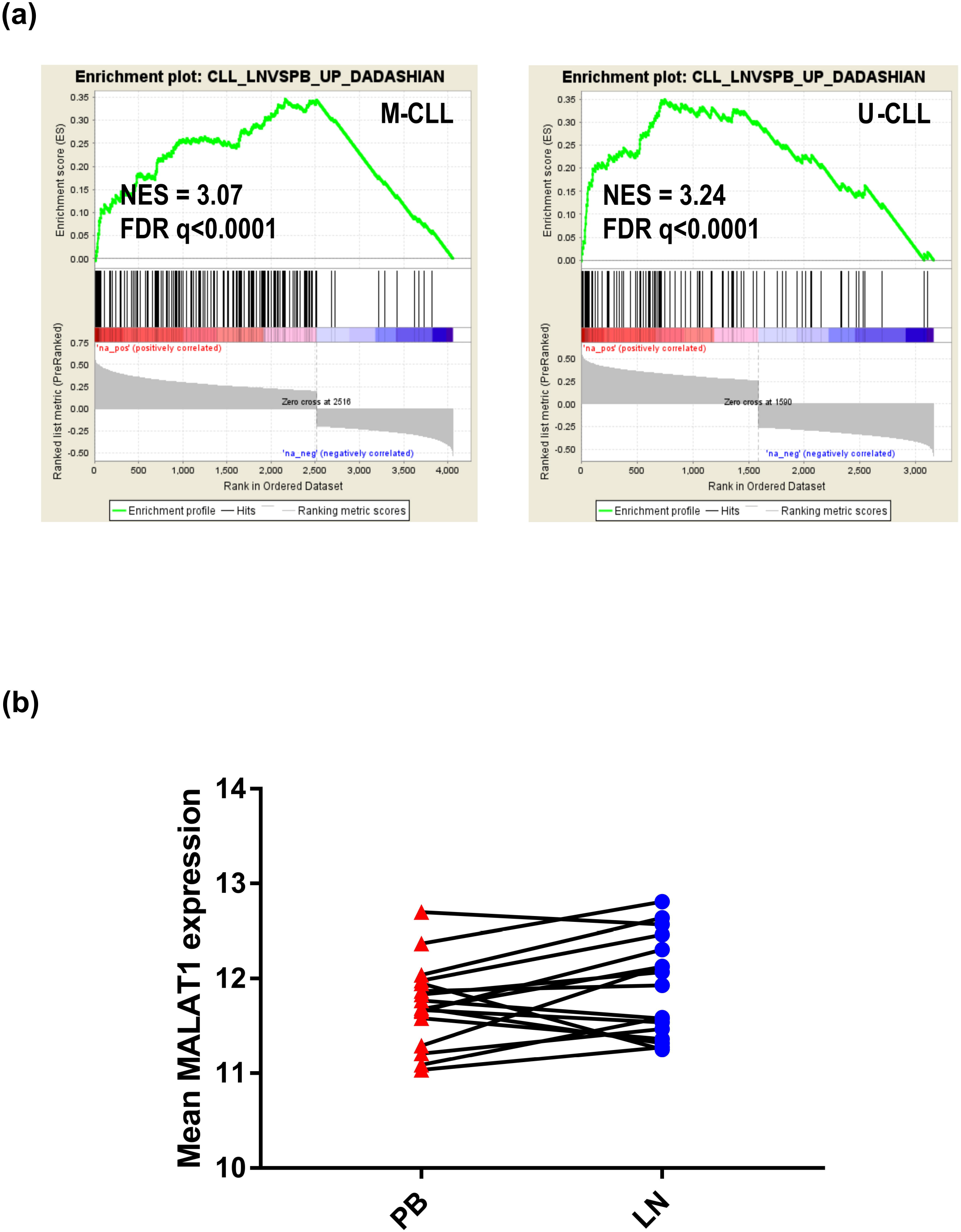
*MALAT1* levels correlate with many genes that are upregulated in LN compared to PB CLL samples, whereas its expression was comparable in both anatomical compartments. **(a)** GSEA analysis shows that the nodal CLL signature (see material and methods) was significantly enriched in coding genes correlated with *MALAT1* expression in PB CLL samples of M-CLL and U-CLL subtypes. **(b)** No significant differences in *MALAT1* expression between paired PB and LN tissues (GSE21029) were observed.

### MALAT1 expression in Follicular Lymphoma

Based on the results of *MALAT1* in CLL and its potential association with the LN microenvironment, we further explored the role of *MALAT1* in FL, another indolent B cell lymphoid neoplasm with predominant nodal presentation. *MALAT1* expression levels were analyzed by qRT-PCR in a series of 61 grade 1-3A FL. These patients had been homogenously treated with R-CHOP. Using the *maxstat* algorithm, we identified two groups of FL with high (N=11) and low (N=50) *MALAT1* expression. The clinical, biological, and histological characteristics, including histological grade, were similar in both subgroups (Table S5). FL cases with high *MALAT1* expression had a significantly shorter progression-free survival (PFS) than cases with low expression (P=0.017) (Fig. 4A). However, no significant differences were observed in transformation to DLBCL or OS between cases with low and high expression (P=0.088 and P=0. 0177) (Fig. S6).

**Figure 4.**
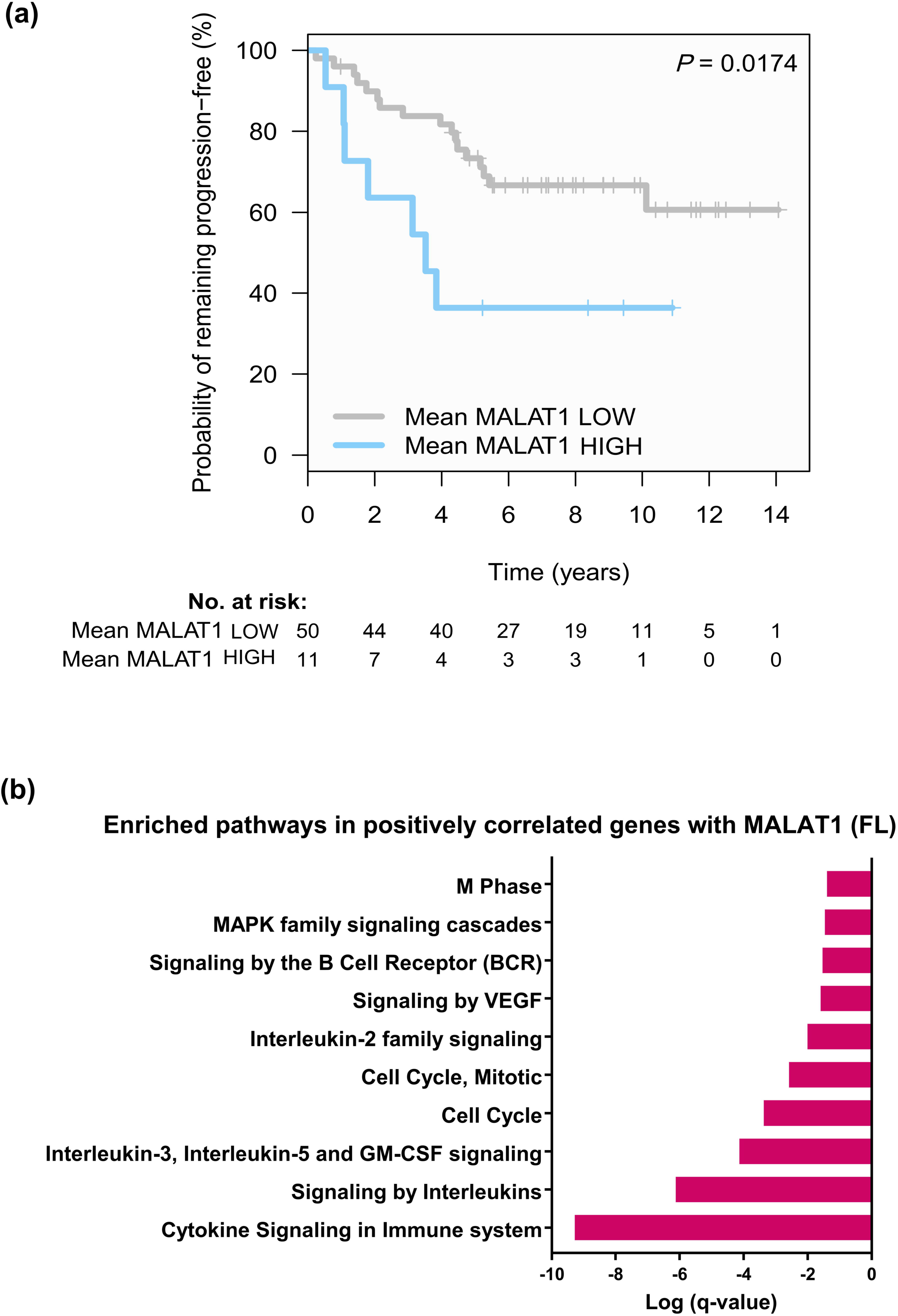
*MALAT1* expression is associated with a more aggressive behavior and is related to pathogenetic pathways in FL. **a**. Kaplan-Meier curves for progression-free survival (PFS) according to *MALAT1* expression in FL. Patients with high *MALAT1* expression had a significantly shorter PFS than those with low levels. **b**. Summary of most relevant significant pathway enrichments among coding genes positively correlated with *MALAT1* in FL samples. Only statistically significant Reactome pathways after multiple comparison correction are shown for compact representation.

To explore the possible biological role of *MALAT1* expression in FL we reanalyzed microarray expression data previously generated on purified FL B-cells^16^. We observed that the coding genes positively correlating with *MALAT1* expression (Table S6) were associated with cell proliferation and other pathways like VEGF, MAPK, interleukin signaling (including some particular pathways related with IL-3 and IL-5) and BCR signaling, also described as involved in FL oncogenesis (Fig. 4B and Table S7)^30,31^. Moreover, among the genes positively correlated with *MALAT1* involved in BCR signaling we noticed *PIK3CD* that it has been previously described as predictor of poor prognosis in FL^32^.

Finally, we performed a comparison of all genes involved in pathways significantly enriched in FL and compared to those found in CLL (considering together U-CLL and M-CLL gene lists). A large proportion of those genes were exclusive for each lymphoma type (88% in FL and 99% in CLL) (Fig. S7 and Table S8). Concordantly, cell cycle-related or MAPK pathways that were initially observed in both neoplasms involved different sets of *MALAT1* positively correlated genes (Tables S9 and Table S10). On the other hand, a total of 12 genes were found in common in the comparative analysis, including several involved in cytokine and interleukin signaling (CAPZA1), cell cycle (GAR1), or both (HSP90AA1, LMNB1 and LYN)(Table S11).

## Discussion

In this report, we provide novel insights into the clinical and biological role of *MALAT1* in indolent B cell neoplasms. Our study revealed that *MALAT1* upregulation was associated with a detrimental clinical behavior in the different entities studied, associated with a shorter TTT in CLL or shorter PFS in FL. Interestingly, *MALAT1* upregulation was a prognostic factor in CLL independently of the IGHV mutational status, epigenetic subgroups, Binet stage or genetic alterations. In FL, *MALAT1* overexpression was also associated with shorter progression-free survival although the clinical and biological features of patients with low and high expression were similar.

We also studied the potential causes and consequences of *MALAT1* upregulation that might justify its clinical behavior in indolent B cell neoplasms. In CLL, *MALAT1* mutations were rare and not related to the expression of the gene. This finding is similar to those in other neoplasms such as bladder cancer, head and neck squamous cell cancer, and lung adenocarcinomas, in which single nucleotides variants and indels were found, although considered passenger events caused by a transcription-associated mutational process^33,34^.

Therefore, other mechanisms should be related with *MALAT1* overexpression and, in fact, its promoter has been described in solid tumor models to be regulated by transcription factors as *HIF1α* and *STAT3*, which are induced by microenvironmental factors as hypoxia and cytokines/interleukins, respectively^35–37^. In CLL, microenvironment stimulation occurs in the LN and is a key process for activation, proliferation and survival of these tumoral B cells^38^. Interestingly, our guilt-by-association analysis on the coding genes correlated with *MALAT1* in PB CLL samples revealed significant enrichments in a gene signature highly expressed in nodal CLL^20^, as well as for interleukin-related signatures. On top of that, we found that *MALAT1* levels remain similar in PB compared to LN paired CLL samples, supporting that the clinical value of *MALAT1* expression found in PB is mirroring expression differences in LN associated with the degree of the microenvironment stimulation.

In FL, our study also revealed biological insights that seem to explain the clinical behavior of *MALAT1* upregulation in these neoplasms. In these tumors, coding genes correlated with *MALAT1* expression were enriched in cell cycle-related processes as well as pathways related to cell proliferation, migration and angiogenesis, such as MAPK and VEGF pathways, which have been previously linked to *MALAT1* function in other tumor models^39^. We also found that cases with high *MALAT1* expression were significantly associated with high levels of genes involved in BCR signaling and interleukin signaling, such as *LYN* and *PIK3CD*, the expression of which promote cell growth and have been associated with poor prognosis in FL^32,40^. In this regard, these are relevant associations supporting our results about the clinical impact of *MALAT1*, even though our series included a relative limited number of cases.

Finally, the comparison of *MALAT1* associated pathways and the involved genes showed a low degree of overlapping in the corresponding gene signatures, even in cell cycle-related or MAPK pathways that were initially observed as enriched in both neoplasms. These results suggest that *MALAT1* could be contributing to the regulation of different transcriptional gene sets in both lymphomas. In spite of these differences at the gene level, we also observed that processes as cell proliferation and MAPK pathways were potentially commonly affected by *MALAT1* upregulation, and thus could explain its common clinical impact in both neoplasms.

In summary, these findings highlight that *MALAT1* overexpression plays a role in the pathobiology and clinical behavior of indolent B cell neoplasms and related with a more aggressive behavior of patients with higher *MALAT1* levels. Particularly in CLL, *MALAT1* could serve as a clinical biomarker that seems to be a surrogate marker of the degree of stimuli the CLL cells receive from the microenvironment. Therefore, *MALAT1* expression could be taken in account in further studies as complementary to other known prognostic factors to improve the clinical management of these patients.

## Data Availability

All data produced in the present study are available upon reasonable request to the authors.

## Statements & Declarations

## Acknowledgements

This research was funded by Ministerio de Ciencia e Innovación (MCI), grant numbers RTI2018-094274-B-I00 to EC.; Fundació La Marato de TV3 (“projecte finançat per Fundació La Marató de TV3”) 201920-30 to LH. Suport Grups de Recerca AGAUR 2014-SGR-795 and 2017-SGR-736 of the Generalitat de Catalunya to EC and JIM-S., respectively. AGAUR 2018 FIB00696, Generalitat de Catalunya to EMF-G. EC and JIM-S are Academia Researchers of the “Institució Catalana de Recerca i Estudis Avançats” (ICREA) of the Generalitat de Catalunya. We are indebted to the Genomics core facility of the Institut d’Investigacions Biomèdiques August Pi I Sunyer (IDIBAPS).

## Authors’ contributions

Conceived and designed the Study: Lluís Hernández and Elías Campo. Collected and processed clinical/expression data from ICGC, Hospital Clinic de Barcelona and GEO: Elena María Fernández-Garnacho, Ferran Nadeu, Pablo Mozas, Andrea Rivero, Julio Delgado, Eva Giné, Armando López-Guillermo, Martí Duran-Ferrer, Itziar Salaverria, Cristina López, Sílvia Beà, Santiago Demajo, Pedro Jares, Xose S Puente and Lluís Hernández; Processed FFPE material from FL samples for RNA purification: Silvia Martín. Analyzed the RNAseq/microarrays/qRT-PCR data and performed the statistical analysis: Elena María Fernández-Garnacho, Ferran Nadeu, Pablo Mozas, Andrea Rivero and Lluís Hernández wrote the manuscript: Elena María Fernández-Garnacho, Lluís Hernández, Elías Campo and José Ignacio Martín-Subero. All authors read and approved the final manuscript.

## Data availability

The data that support the findings of this study are available from the corresponding author upon reasonable request.

## Competing interests

The authors declare that they have no competing interests.

## Ethics approval and consent to participate

Samples from FL patients were obtained from the Hematopathology collection of the Biobank of the Hospital Clínic de Barcelona-IDIBAPS (Spain). RNAseq data and full clinical annotations from CLL patients were obtained from the Spanish ICGC consortium, which had an approval of the Institutional Review Board of Hospital Clinic de Barcelona, in accordance with national regulations and the Declaration of Helsinki. All patients provided written informed consent. The remaining data from other samples were obtained from GEO microarray public repository including previously published works with their particular ethical compliances stated in the original articles.

## Supplementary Figure Legends

**Fig.S1. (a)** Optimal cut-off established by *maxstat* algorithm regarding *MALAT1* expression and TTT in CLL. **(b)** Optimal cut-off established by *maxstat* algorithm regarding *MALAT1* expression and OS in CLL.

**Fig.S2. (a)** Binet A CLL cases with high *MALAT1* expression had a significantly shorter TTT than those with low levels. **(b)** No difference in OS was detected in all CLL cases with high or low *MALAT1* expression neither using the same cutoff defining significant differences for TTT (see Suppl. Fig1b) nor the optimal for OS (data not shown). **(c)** TTT and *MALAT1* expression levels in IGHV-mutated and unmutated Binet A CLL cases. **(d)** TTT and *MALAT1* expression levels in Binet A CLL epigenetic subtypes.

**Fig.S3. (a)** Similar *MALAT1* levels in CLL cases with a different number of genetic alterations. **(b)** No significant differences in *MALAT1* expression comparing CLL cases with (mutated) or without (wild-type) individual gene alterations after correction for multiple comparisons. **(c)** No significant differences in *MALAT1* expression comparing CLL cases with (altered) or without (wild-type) individual chromosomal alterations after correction for multiple comparisons.

**Fig.S4. (a)** Boxplot showing the distribution of the values of the three epiCMIT indexes in the three epigenetic groups of CLL cases analyzed according to the *MALAT1* expression categories. No significant differences were found in any comparison. **(b)** Scatterplots showing the linear correlation analyses between the three epiCMIT indexes and *MALAT1* expression values, in the three CLL epigenetic subtypes. None of these correlations were statistically significant.

**Fig.S5**. Summary of most relevant significant pathway enrichments found using Metascape tool involving signatures previously related to CLL pathogenesis and poor prognosis. Separated analyses were performed involving coding genes positively (top panel) and negatively (bottom panel) correlated with *MALAT1* expression in the different epigenetic CLL subtypes. Only statistically significant Reactome pathways after multiple comparison correction are shown for compact representation.

**Fig.S6. (a)** Kaplan-Meier curve showing the lack of significant differences in OS of FL cases regarding *MALAT1* expression. **(b)** Cumulative incidence curves showing the lack of significant differences regarding *MALAT1* expression and the risk of transformation.

**Fig.S7**. Venn diagram showing the overlapping between FL and CLL of *MALAT1*-correlated genes in pathways found significantly enriched in separated analysis.

## Notes

### Competing Interest Statement

The authors have declared no competing interest.

### Funding Statement

This research was funded by Ministerio de Ciencia e Innovacion (MCI), grant numbers RTI2018-094274-B-I00 to EC.; Fundacio La Marato de TV3 (projecte financat per Fundacio La Marato de TV3) 201920-30 to LH. Suport Grups de Recerca AGAUR 2014-SGR-795 and 2017-SGR-736 of the Generalitat de Catalunya to EC and JIM-S., respectively. AGAUR 2018 FIB00696, Generalitat de Catalunya to EMF-G. EC and JIM-S are Academia Researchers of the "Institucio Catalana de Recerca i Estudis Avancats" (ICREA) of the Generalitat de Catalunya.

### Author Declarations

Ethics committee/IRB of Hospital Clinic de Barcelona gave ethical approval for this work as FL samples were obtained from the Hematopathology collection of the Biobank of Hospital Clinic de Barcelona-IDIBAPS and CLL samples data and clinical annotations were obtained from the project of the Spanish ICGC consortium. The remaining data from other samples were obtained from GEO microarray public repository including previously published works with their particular ethical compliances stated in the original articles.

