## Supplementary figures and images for "*MALAT1* Expression is Associated with Aggressive Behavior in Indolent B-Cell Neoplasms"

### FigS1

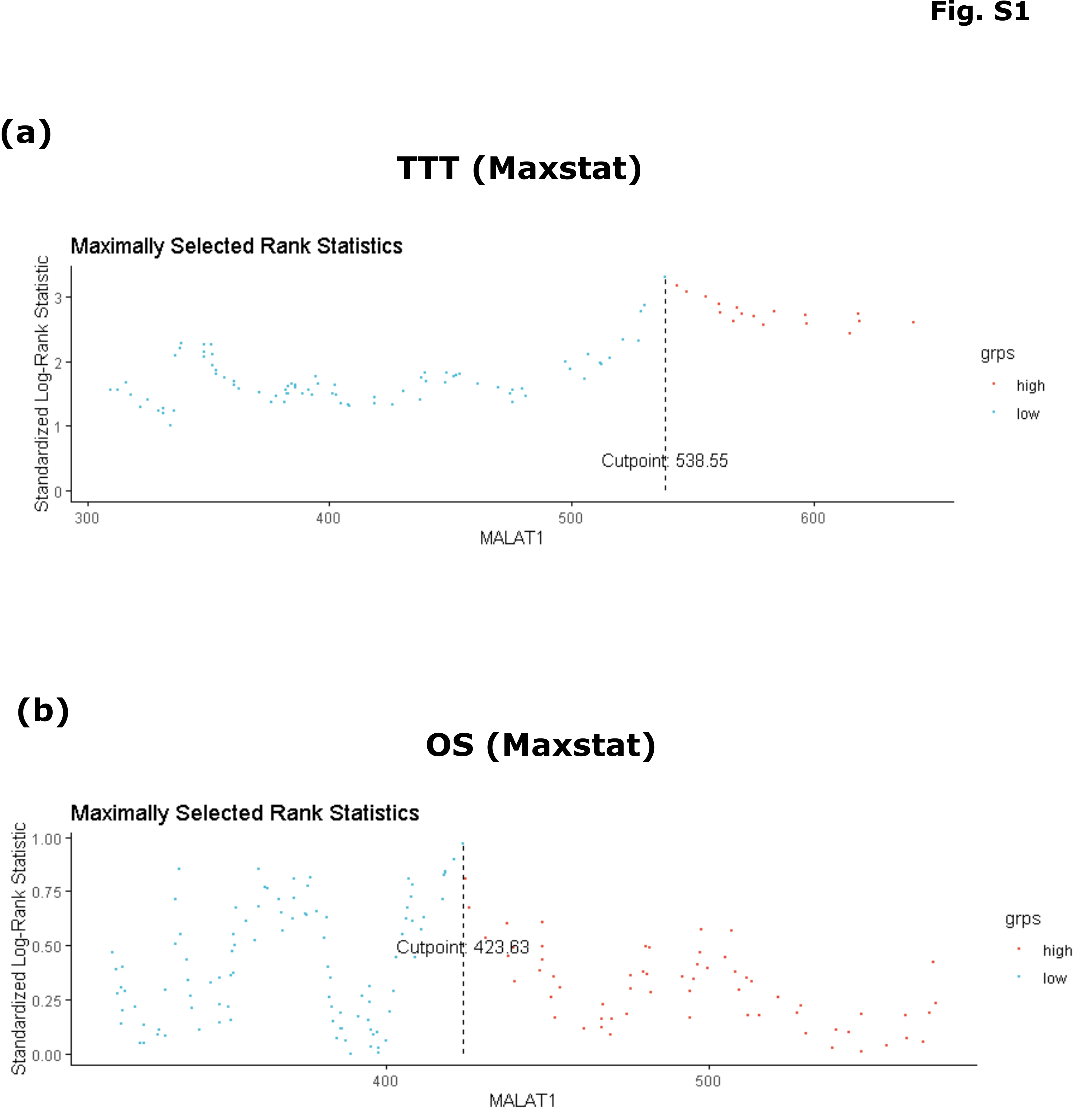

### FigS2

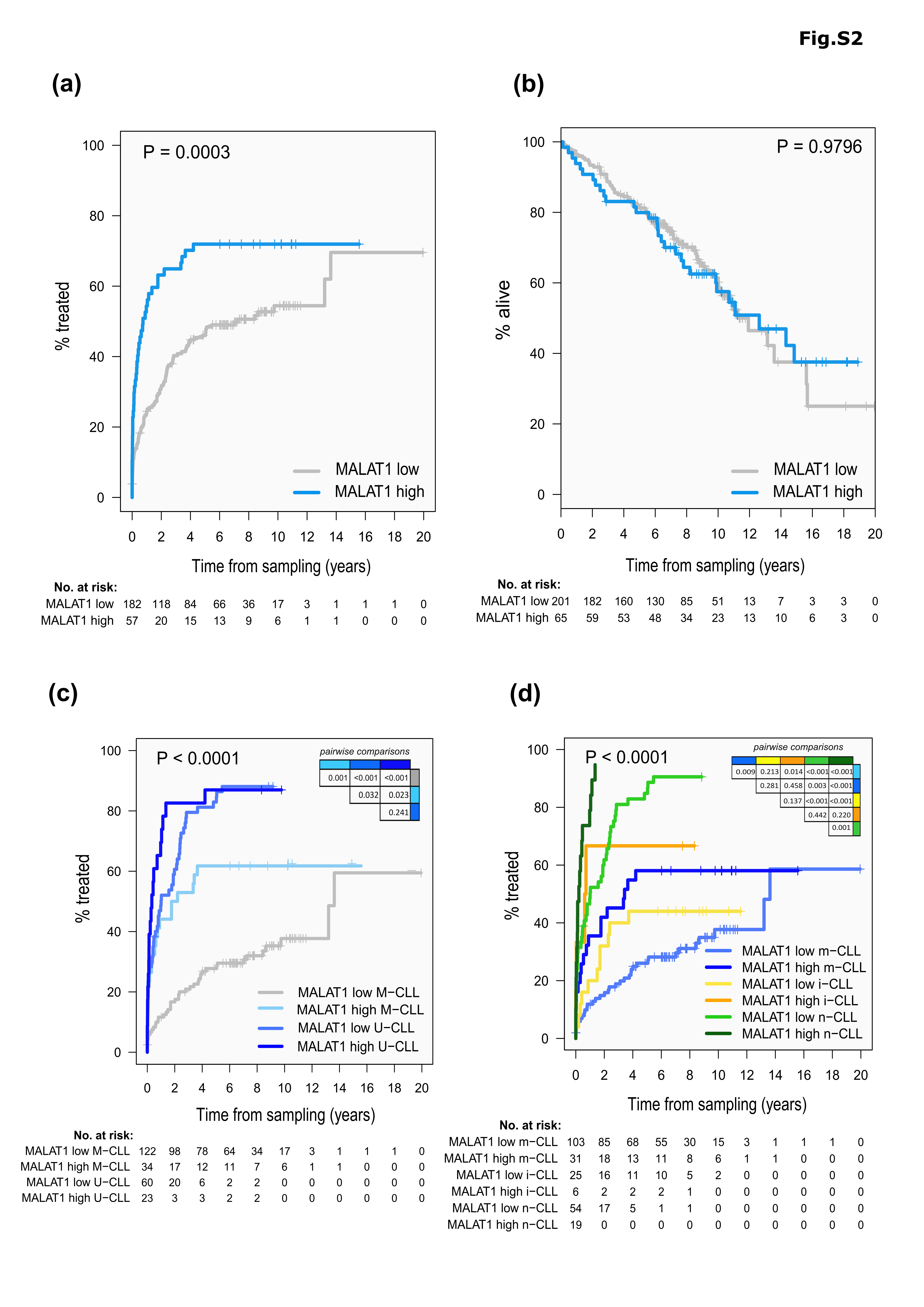

### FigS3

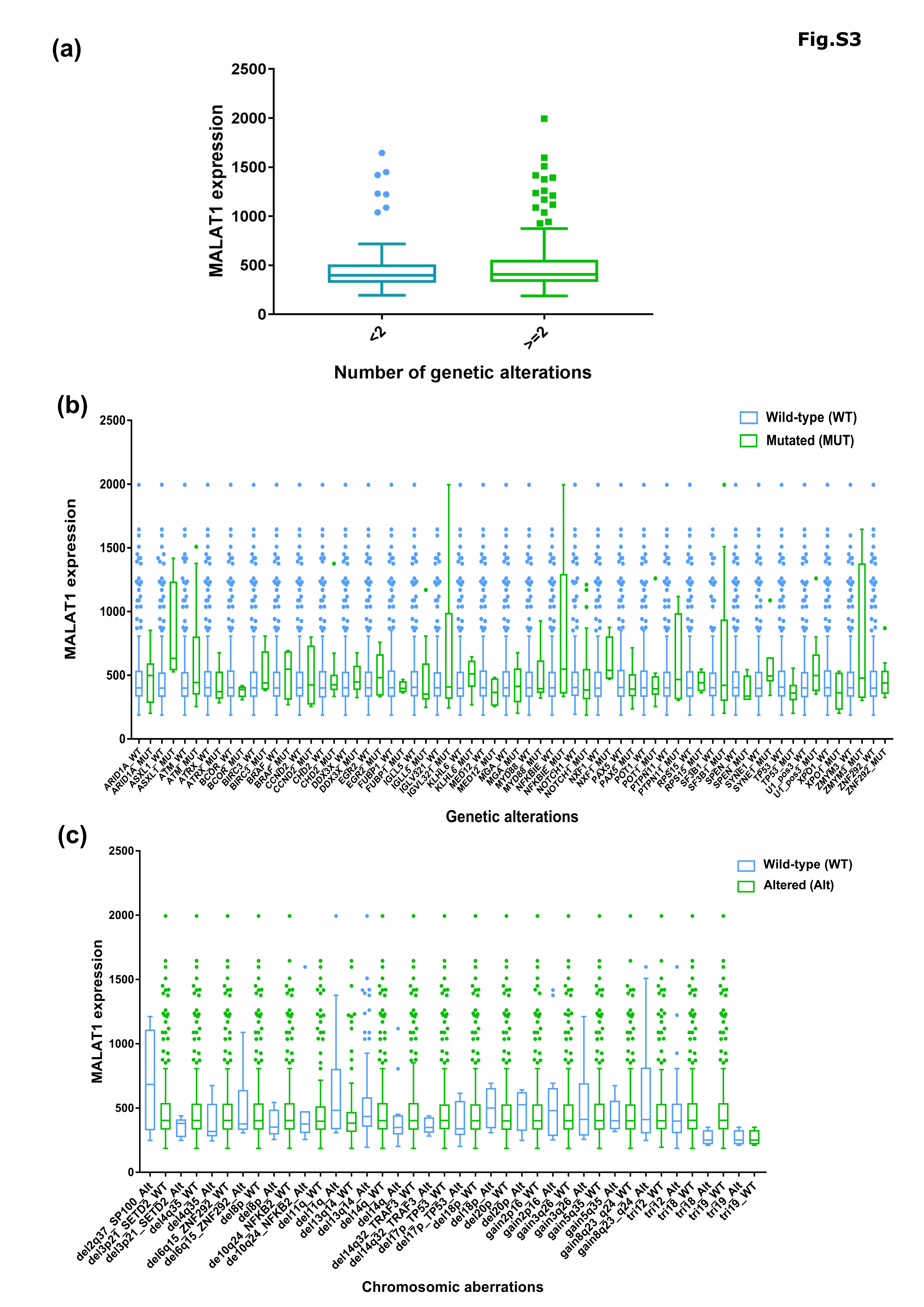

### FigS4

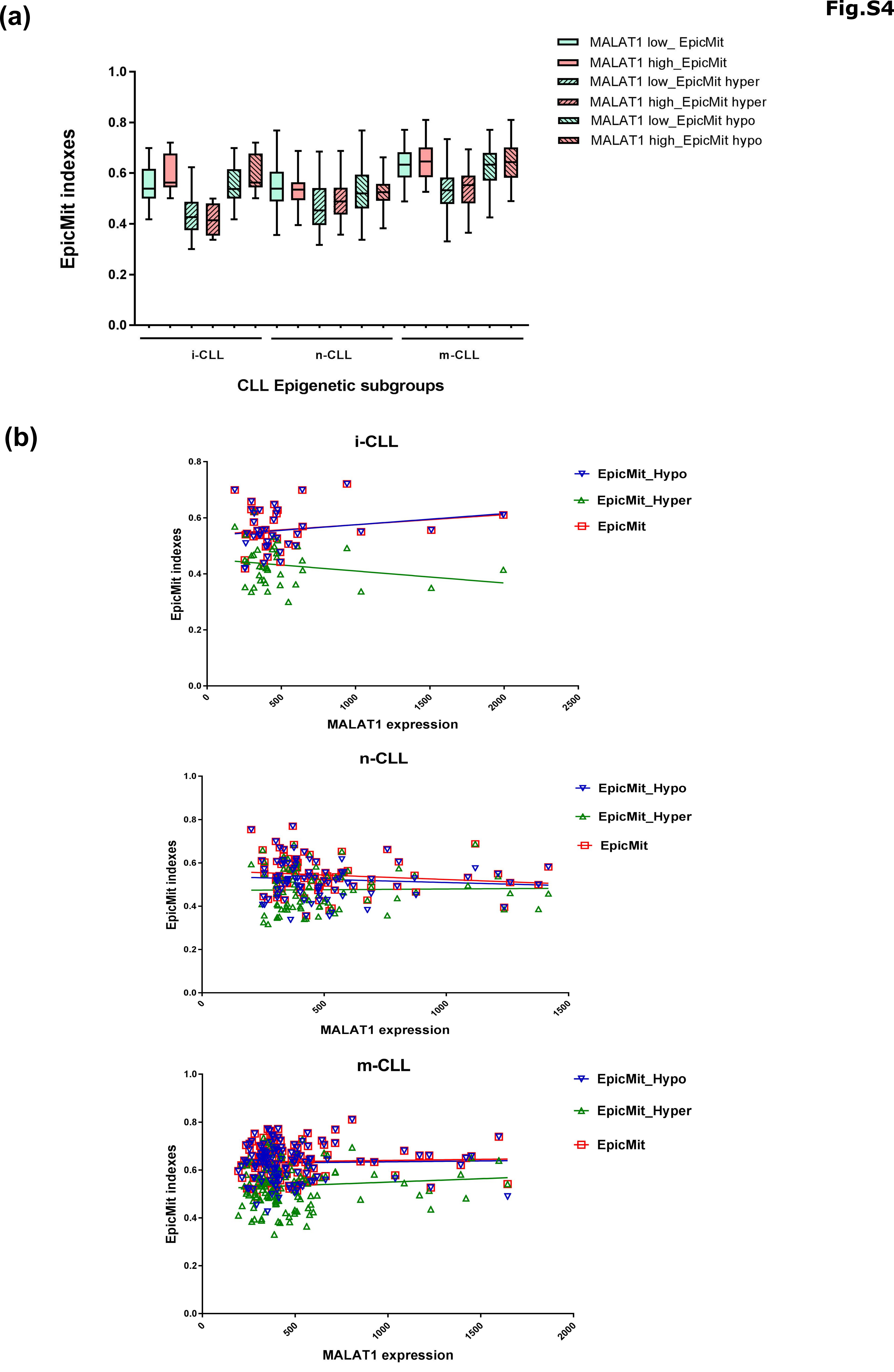

### FigS5

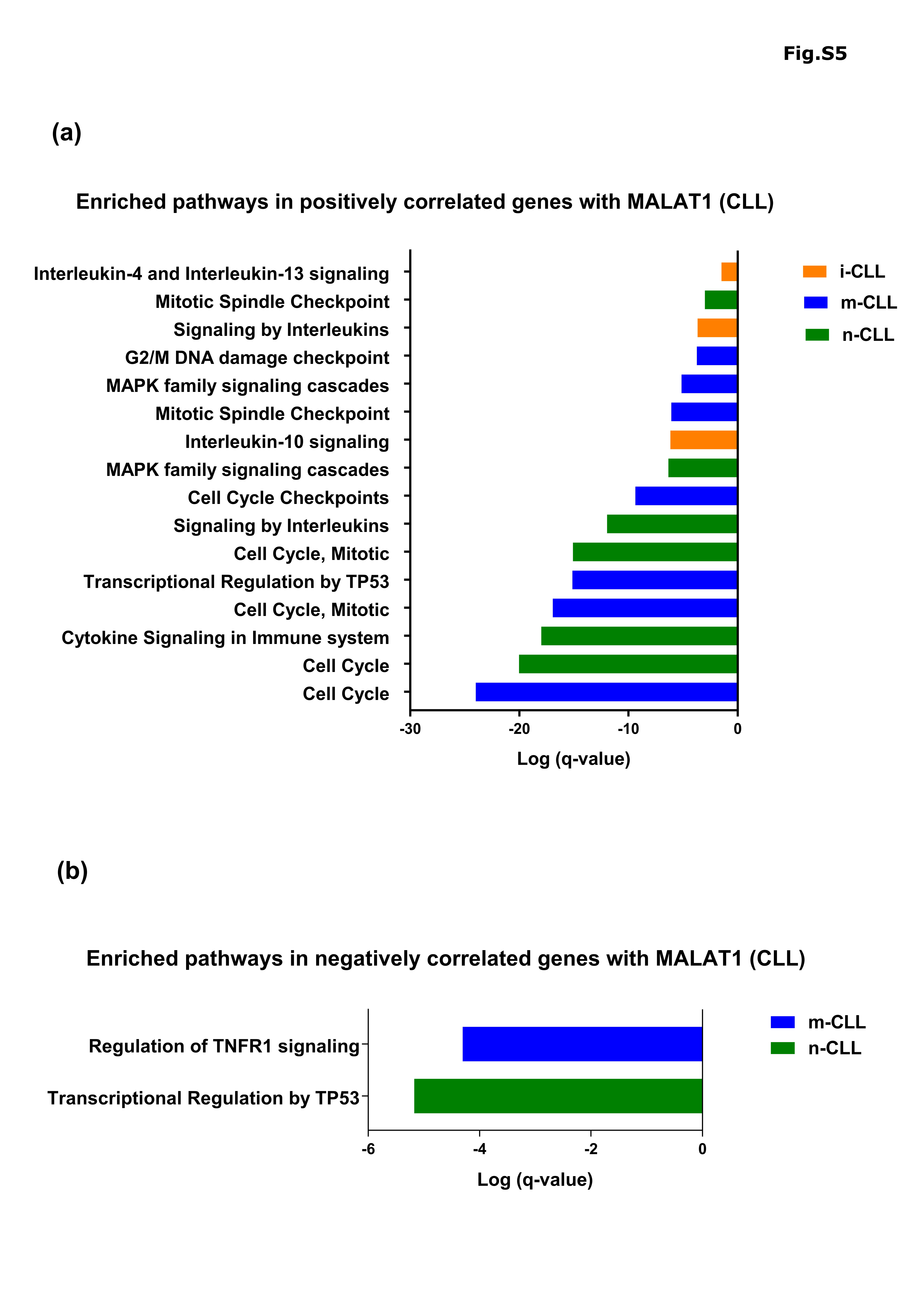

### FigS6

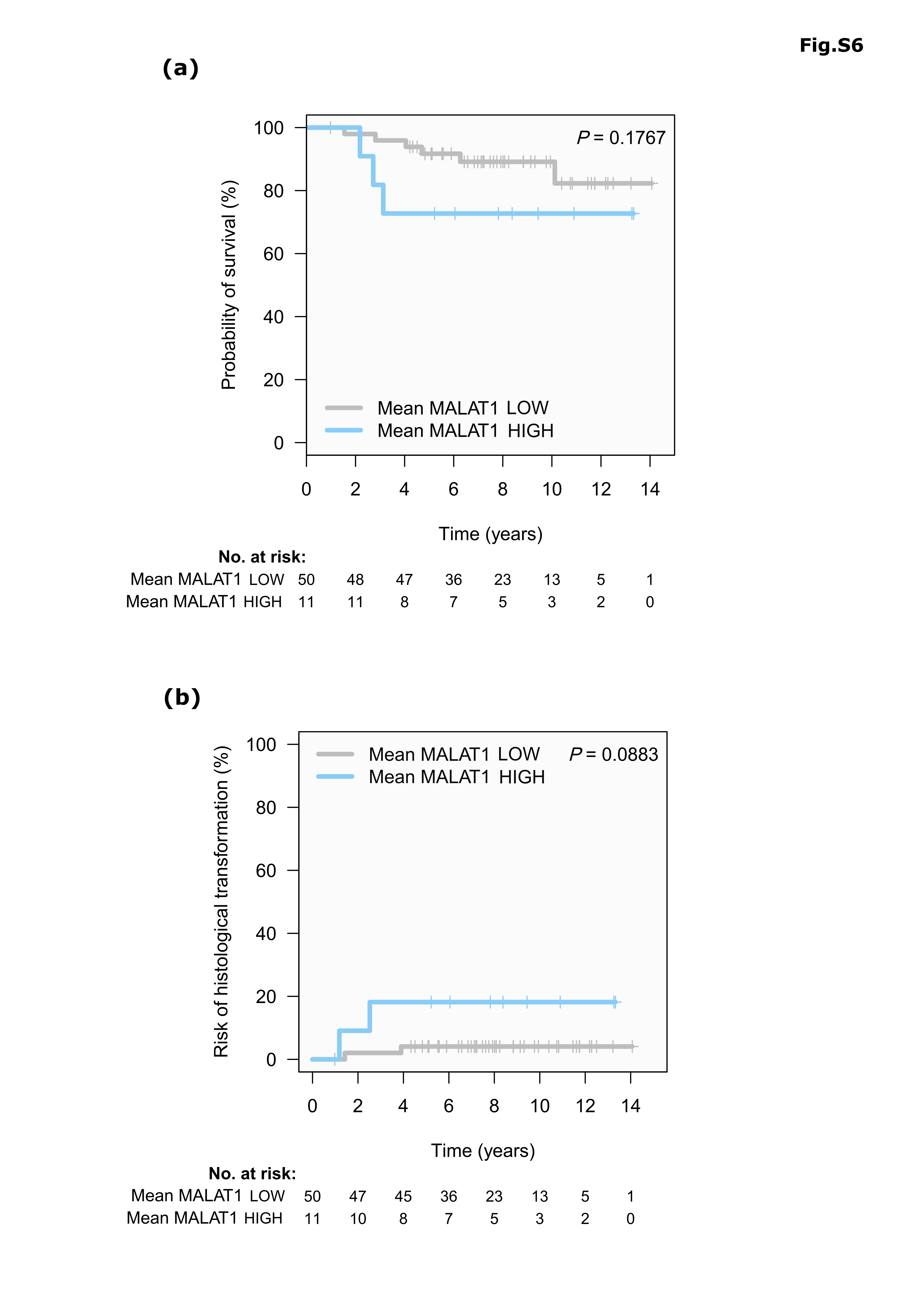

### FigS7

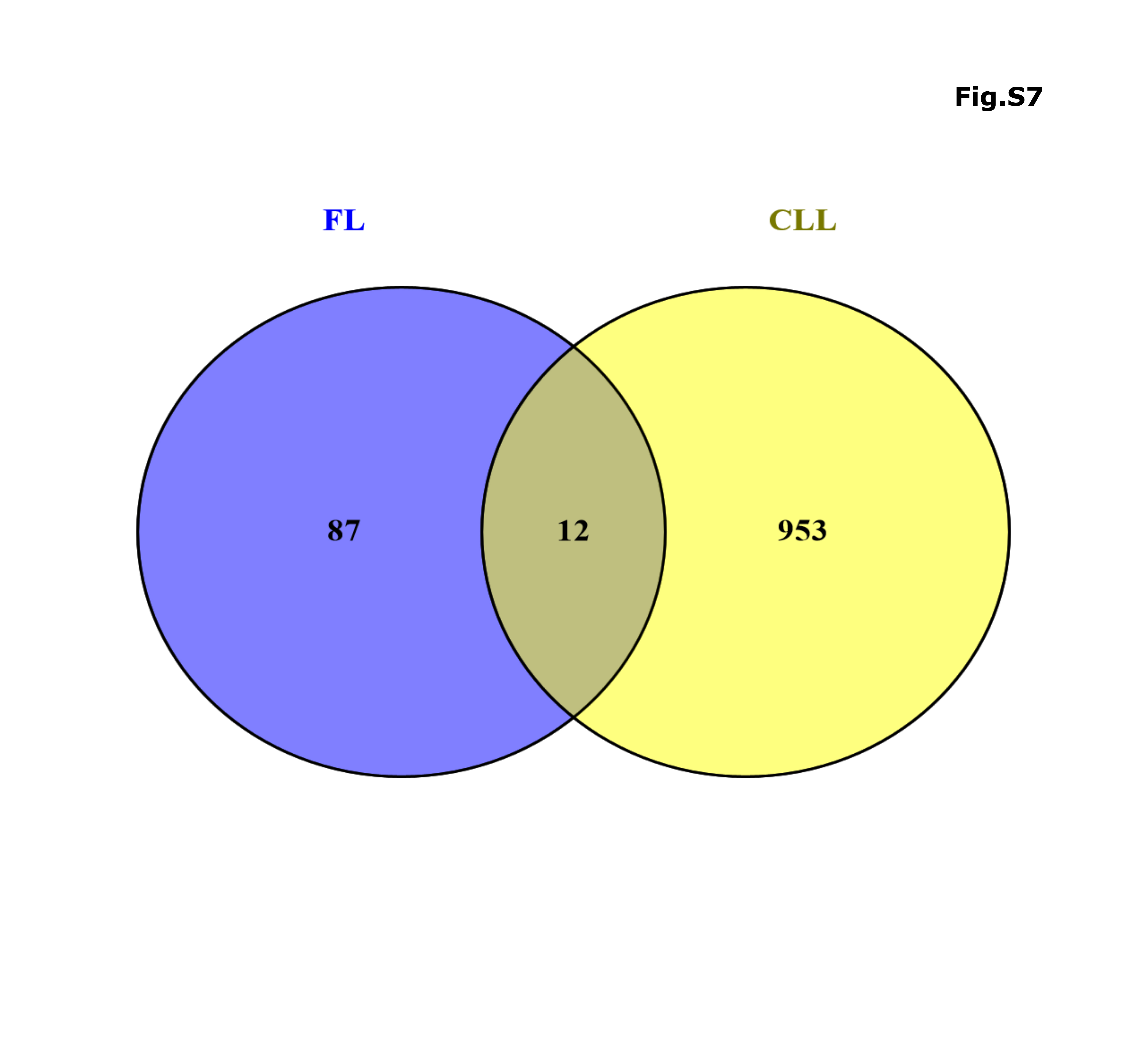
